# Disparities in Vulnerability to Severe Complications from COVID-19 in the United States

**DOI:** 10.1101/2020.05.28.20115899

**Authors:** Emily E. Wiemers, Scott Abrahams, Marwa AlFakhri, V. Joseph Hotz, Robert F. Schoeni, Judith A. Seltzer

## Abstract

This paper provides the first nationally representative estimates of vulnerability to severe complications from COVID-19 overall and across race-ethnicity and socioeconomic status. We use the Panel Study of Income Dynamics (PSID) to examine the prevalence of specific health conditions associated with complications from COVID-19 and to calculate, for each individual, an index of the risk of severe complications from respiratory infections developed by DeCaprio et al. (2020). We show large disparities across race-ethnicity and socioeconomic status in the prevalence of conditions which are associated with the risk of severe complications from COVID-19. Moreover, we show that these disparities emerge early in life, prior to age 65, leading to higher vulnerability to such complications. While vulnerability is highest among older adults regardless of their race-ethnicity or socioeconomic status, our results suggest particular attention should also be given to the risk of adverse outcomes in midlife for non-Hispanic Blacks, adults with a high school degree or less, and low-income Americans.

---

The presence of preexisting health conditions increases vulnerability to severe complications from COVID-19 (1). The CDC reports that over 90% of hospitalized patients have at least one preexisting health condition (2). In hospitals in New York City, Long Island, and Westchester County, NY, 94% of patients hospitalized had at least one preexisting condition, 88% had more than one, and the median number was four (3). The most common conditions among patients hospitalized for COVID-19 are hypertension (3–13), diabetes, cardiovascular disease, kidney disease, and obesity (3, 6–12). Conditional on hospitalization, those with preexisting conditions suffer more severe outcomes (1, 6, 10-12) and experience higher fatality rates in China and the United States (1, 11, 14).

In the United States, vulnerability to COVID-19 based on preexisting health conditions, collides with long-standing disparities in health and mortality by race-ethnicity and socioeconomic status, especially in midlife (15–17). Recent studies have highlighted these disparities finding that Blacks, and the socioeconomically disadvantaged are more likely to have at least one health condition that makes them vulnerable to COVID-19 (18, 19). Indeed, COVID-19 hospitalization rates are higher for Blacks, and Hispanics while Whites are underrepresented relative to their population percentages (20, 21). Differences in mortality rates by race-ethnicity are staggering: in Washington D.C., African Americans make up 45% of the population yet suffer 76% of deaths (22). Yet what is lacking is a means of translating the well-known disparities in a multiplicity of health conditions into predictions about how the likelihood of having serious complications from COVID-19 varies across race-ethnicity and socioeconomic status. As the pandemic continues, the inability to translate health disparities into vulnerability to COVID-19 hampers our ability to be forward-looking about differential impact of the virus.

This paper leverages a recent machine learning model of the likelihood of hospitalization for several respiratory conditions (23). We apply the model to preexisting health conditions reported in the Panel Study of Income Dynamics (PSID) which allows us to form the first population-representative estimates of vulnerability to in-patient hospitalization for respiratory conditions for the U.S. and how vulnerability varies by education, income, and race-ethnicity. As the vulnerability index is itself a function of preexisting health conditions, we also examine the distribution of such conditions across demographic groups.

We find that disparities in preexisting health conditions across race-ethnicity and socioeconomic status translate into large gaps in vulnerability. Adults with a high school education or less are 1.6 times as likely as those with a college degree to face severe complications and Americans in the lowest income quartile face 2.7 times the risk compared to those in the highest quartile. These gaps are due to preexisting health conditions that emerge earlier in life for less-educated and low-income adults. We also find large gaps in vulnerability appearing in midlife for non-Hispanic Blacks, whereas Hispanics face lower risks based on preexisting health conditions and age. We show that to understand disparities in vulnerability to severe complications from COVID-19, it is crucial to describe how disparities in the age pattern of specific preexisting conditions and the presence of multiple conditions would account for disparities in the incidence of severe complications from the COVID-19 pandemic in the U.S.

We also show that while disparities in preexisting health do predict disparities in complications from COVID-19, our predictions do not fully track the disparities in observed hospitalization from the virus. The differences in our predictions and actual hospitalizations suggests that there are differences by race-ethnicity and socioeconomic status in exposure to the coronavirus, the likelihood of infection given exposure, and in the complications from infections that go beyond the disparities in preexisting health. Disparities in exposure based on residential and occupational characteristics have been documented during the COVID-19 pandemic (6, 7). The persistence of race-ethnic differences in hospitalization rates for COVID-19 after controlling for comorbidities suggests there may be disparities in timely access to care (24). However, understanding variation in vulnerability based on preexisting health conditions is crucial for predicting the likely disparities in the effect of the disease and for assessing the role of factors beyond health conditions. For these tasks, population-representative estimates are essential. While we focus on disparities in preexisting health conditions and their implications for vulnerability to severe complications, our work is a complement to studies of disparities in exposure to the coronavirus (25–27) and in inequities in health care (24, 28, 29).

## MATERIALS AND METHODS

In this section we discuss the data and methods used to analyze the preexisting health conditions that the CDC and the developing literature indicate are risk factors for severe illness from COVID-19. We begin with a discussion of the Panel Study of Income Dynamics (PSID) data that we use to determine the distribution of and disparities in risk factors in the adult U.S. population. We also describe the index of relative vulnerability to severe illness from respiratory infections developed by ClosedLoop.ai (23), which is a function of preexisting conditions, recent hospitalization, and age. We discuss the construction of the index and how we apply it to nationally representative data.

### Panel Study of Income Dynamics

The PSID is a national longitudinal survey that has interviewed the original sample members and their descendants since 1968. The original sample included oversamples of Black and low-income families to facilitate the study of poverty. With the addition in 1997 of a sample of immigrants who arrived in the United States after 1968 plus the addition in 2017 of a sample of immigrants who arrived after 1997, the 2017 wave includes 26,455 individuals representative of the national population when weighted (30). We use the 2017 data on adults ages 25 and older, a sample of 13,150 household heads and spouses (or cohabiting partners)^1^ who are not missing data on age, race-ethnicity, educational attainment, household income, or preexisting health conditions.^2^ Throughout our analyses, we apply the 2017 PSID cross-sectional weights.

#### Demographics and Socioeconomic Status

Race-ethnicity is classified as Hispanic, non-Hispanic Black, non-Hispanic White, and non-Hispanic other. Because of sample size we do not report separate estimates for individuals who are non-Hispanic other race. Education is categorized as no more than 12 years (high school/GED or less), 13-15 years (some college), and at least 16 years (BA degree or more). Income is measured at the household level. We classify each adult according to the income quartile of their household.^3^

The population characteristics of our sample are displayed in Table 1. We show the distributions of gender, race-ethnicity, educational attainment, and household income quartile overall and for three age groups (25-44, 45-64 and 65+). We also show the distribution of age across each race-ethnic, educational attainment, and household income category. In Appendix A, we compare the PSID data to that of the Current Population Survey (CPS) from the 2017 Annual Social and Economic Supplement (ASEC). Overall, the samples produce almost identical distributions.^4^ Prior work has shown that estimates of income from the PSID match well with those from the March CPS (31).

**Table 1.** Percent of Demographic and SES Groups by Age across Age Categories & Mean Age

|  | Percent across Age Categories |  |  | Percent within Age Categories |  |  | Percent,<br>All Ages<br>(25+) | Mean<br>Age | Sample<br>sizes |
| --- | --- | --- | --- | --- | --- | --- | --- | --- | --- |
|  | 25-44 | 45-64 | 65+ | 25-44 | 45-64 | 65+ |  |  |  |
| Overall |  |  |  | 38 | 38 | 24 |  | 51.9 | 13,150 |
| Gender |  |  |  |  |  |  |  |  |  |
| Female | 37 | 38 | 26 | 51 | 52 | 55 | 52 | 52.4 | 7,140 |
| Male | 39 | 38 | 23 | 49 | 48 | 45 | 48 | 51.5** | 6,010 |
| Race and Ethncity |  |  |  |  |  |  |  |  |  |
| NH White | 33 | 38 | 28 | 64 | 72 | 83 | 71 | 53.6 | 7,082 |
| NH Black | 42 | 41 | 17 | 12 | 12 | 7 | 11 | 49.2*** | 4,158 |
| Hispanic | 52 | 36 | 13 | 18 | 12 | 7 | 13 | 46.5*** | 1,464 |
| NH Other | 52 | 32 | 15 | 7 | 4 | 3 | 5 | 47.4*** | 446 |
| Education |  |  |  |  |  |  |  |  |  |
| HS or less | 32 | 40 | 28 | 32 | 41 | 44 | 38 | 54.3*** | 5,262 |
| Some college | 39 | 38 | 23 | 25 | 24 | 22 | 24 | 51.2** | 3,482 |
| BA or more | 43 | 36 | 22 | 43 | 36 | 34 | 38 | 50.1 | 4,406 |
| Household Income |  |  |  |  |  |  |  |  |  |
| Bottom Quartile | 31 | 33 | 37 | 15 | 16 | 27 | 18 | 56.6*** | 2,550 |
| Second Quartile | 40 | 31 | 29 | 24 | 18 | 27 | 23 | 52.3*** | 3,166 |
| Third Quartile | 41 | 36 | 23 | 30 | 26 | 26 | 28 | 50.5 | 3,660 |
| Top Quartile | 37 | 48 | 15 | 31 | 40 | 20 | 32 | 50.3 | 3,774 |
| Sample sizes | 6,808 | 4,392 | 1,950 |  |  |  |  |  |  |

Data Source: 2017 Wave, PSID. Sample: PSID heads and spouses, 25 years and older. Weights: PSID individual cross-section weights.
**Notes:** Asterisks (\*) are for tests of age of a subgroup being significantly different from base group, where: \* P-value $\leq 0.10$ ; \*\* P-value $\leq 0.05$ ; \*\*\* P-value $\leq 0.01$ . Base groups are female, NH White, BA or more, top income quartile.

Differences in the age distribution across demographic and socioeconomic groups matter because the risks from COVID-19 vary with age. Females are one year older, which is consistent with well-documented higher mortality rates for men. Non-Hispanic Whites (NH White) make up 71% of the adult U.S. population, while 11% are Non-Hispanic Blacks (NH Blacks), 13% are Hispanics and 5% are other non-Hispanic groups. NH Whites are the oldest on average (53.6 years old), followed by NH Blacks (49.2), and Hispanics are the youngest at 46.5 years. Among NH Whites, 28% are 65 and older compared to only 17% of NH Blacks, and 13% of Hispanics. With respect to educational attainment, 38% of people have a high school degree or less, 24% have completed some college, and 38% have a BA degree or higher. Due to educational differences across birth cohorts, those with a high school degree or less are older (54.3 years old) than those with some college (51.2) or those with a BA (50.1). Finally, 18% of individuals in the adult population are in the lowest quartile of household income, while 23%, 28% and 32% are in the second, third and top quartiles, respectively. Again, we see age differences across these quartiles, with those in the lowest quartile being older (56.6 years) compared to the top two quartiles, which average 50.4 years.

#### Self-Reported Health Conditions (Risk Factors)

The PSID includes questions about whether a person was ever told by a health professional they have certain health conditions. Many of these conditions are listed by the CDC as risk factors for becoming severely ill from infection of COVID-19. The health conditions (risk factors) in the PSID include asthma, diabetes, heart disease, heart attack, hypertension, lung disease, neurological conditions, cancer, stroke, and kidney disease. In addition, the PSID asks respondents whether they were hospitalized in the previous year. Past hospitalization is a risk factor in the DeCaprio et al. model described below. Finally, height and weight are reported, which we use to compute each respondent’s BMI. We then create an indicator for severe obesity (BMI ≥ 40). Appendix B describes how estimates of the prevalence of self-reported health conditions in the PSID compare to those in other data sources. Appendix Table B-1 shows the question wording for all self-reported health conditions in the PSID.

### Relative Vulnerability Index

According to the CDC, the risk of severe complications from COVID-19 resulting in hospitalization increases with a number of preexisting health conditions, i.e., risk factors or comorbidities, and with age, but the size of the effect of each risk factor is currently unknown (2). To circumvent the lack of current data on these effects, DeCaprio et al. (23) assessed the risk of hospitalization for respiratory infections available in existing medical claims data. In particular, they looked at (in-patient) hospitalizations associated with: acute respiratory distress syndrome, pneumonia (other than caused by tuberculosis), influenza, acute bronchitis and other upper respiratory infections. DeCaprio et al. used medical claims data on hospitalizations for Medicare recipients – using the Center for Medicare and Medicaid Services Limited Data Set, which contains a 5% sample of the Medicare population – and from Healthfirst, a New York, non-Medicare population insurer. They mapped in-patient hospitalizations from these respiratory infections to individual-level data on 11 preexisting health conditions (risk factors) coded in the Medicare and Healthfirst data, along with patients’ gender, age and whether they had been hospitalized in the last three months.

As described in DeCaprio et al. (23), the authors estimated several alternative versions of their model. In our paper, we use their “Survey Model,” which is based on a logistic regression of incidence of hospitalizations due to the above conditions and is suitable for use with the health conditions available in the PSID.^5^ The model is trained on over 2 million individuals.

The model is of the following form. Let *d*_*i*_ denote a 0/1 indicator of whether individual *i* is hospitalized with a severe respiratory infection. Then,

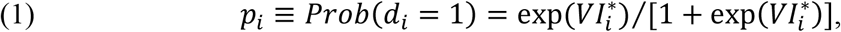

where *d*_*i*_ is a 0/1 indicator for whether *i* is hospitalized and *VI*_*i*_ denotes individual *i*’s *Vulnerability Index score* which is given by the following function:

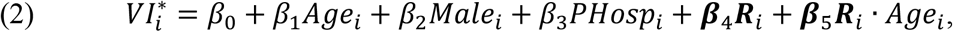

where *Age*_*i*_ is *i*’s current age, *Male*_*i*_ is a 0/1 indicator of whether *i* is a male; *PHosp*_*i*_ is a 0/1 indicator of whether *i* has been recently hospitalized, and ***R***_*i*_ is a vector of 0/1 indicators for whether *i* has particular health conditions, or *risk factors, R*_*ki*_ *k, k* = 1,…, *K*. Given the logistic form of (1), it follows that 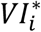 is the *log odds* of severe illness for individual *i* that is a function of their age, gender, recent hospitalization and health-related risk factors. It follows that

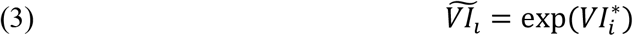

is their *odds ratio* of risk for a severe illness or the *relative vulnerability index* score for individual *i*.

Given the expression for 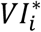 in (2), the base group for the DeCaprio et al. model is a female with no preexisting risk factors and age equal to 0. But, to provide a more meaningful base group, we use the following relative vulnerability index score:

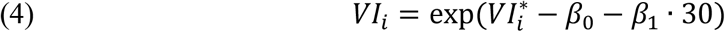

so *VI*_*i*_ provides the odds that individual *i* has of a severe illness relative to that for a 30-year-old female with no risk factors. We report average *VI*_*i*_ by age, race-ethnicity, education, and household income.

In Appendix Table B-2, we display, in the first two columns, respectively, the risk factors in ***R***_*i*_ for the DeCaprio et al. model and estimates of the *β*_*j*_’s for 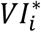 in (2). The third column in Appendix Table B-1 indicates which health risk factors are available in the PSID. As one can see, the PSID does not measure some risk factors included in the DeCaprio et al. model. In our implementation of the vulnerability index with PSID data, we do not attempt to adjust the formula for absent variables; rather, we act as if the members of the PSID sample do not have these conditions. Because of this, the values of the vulnerability index scores for our sample will be biased downwards. As discussed in Appendix B, almost all of the risk factors used in the DeCaprio et al. model that are not available in the PSID, with the exception of liver disease (cirrhosis of the liver), either have very low marginal impacts on the vulnerability index (based on the coefficient estimates for these conditions) or the incidence of the risk factor is very low in the U.S. population.

Finally, to demonstrate how the various risk factors affect the relative vulnerability index scores, we present, in Figure 1, the age profiles of the marginal contributions of each risk factor to relative vulnerability. The marginal contribution at age, *Age*_*i*_, for risk factor, *R*_*ki*_, is given by:

**Figure 1.**
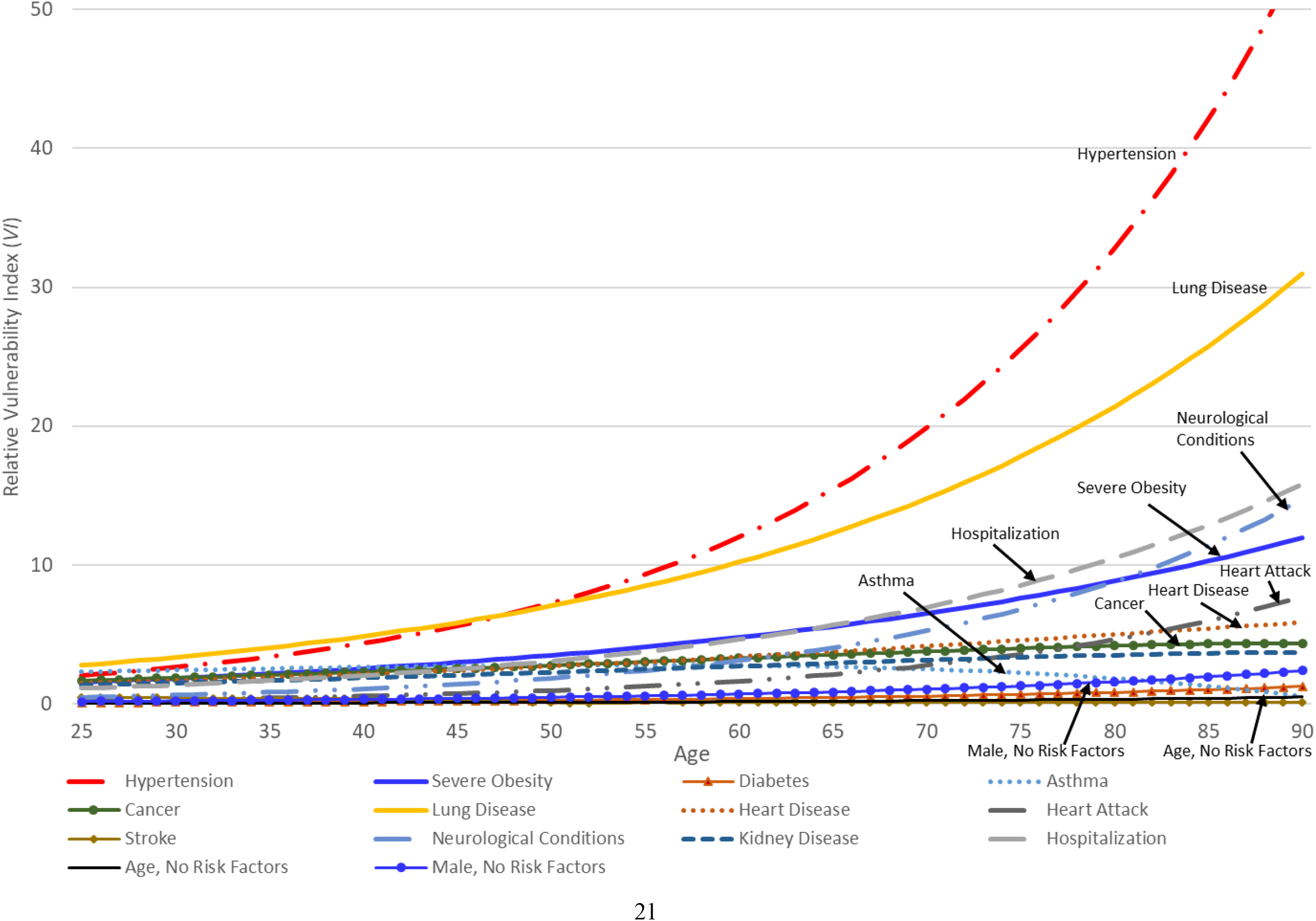
Marginal Contribution of Each Risk Factor Available in PSID to Relative Vulnerability Index (*VI*) by age

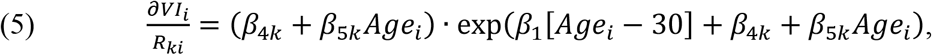

where we evaluate all of the other risk factors, *R*_*mi*_, *m* ≠ *kk*, at 0.

In Figure 1, we draw attention to those risk factors that have the most sizeable contributions to vulnerability at various ages by labeling them in the figure. As one can see from Figure 1, hypertension and lung disease have the largest impacts on the relative vulnerability index at all ages, and both effects rise rapidly with age. Severe obesity – and, to a lesser extent, cancer – also consistently have a large impact on the vulnerability index at all ages. In early adulthood (25–44), asthma contributes a sizeable impact on relative vulnerability, but this risk factor is less important at midlife (45-64) and older ages (65+). With age, one also sees that heart disease and heart attacks contribute to relative vulnerability. At older ages (65+), neurological conditions start to have substantial impacts on relative vulnerability. Finally, recent hospitalizations, which presumably proxy for compromised health, have larger impacts on relative vulnerability as individuals age.

All of the other risk factors have more modest impacts, raising the vulnerability odds of severe disease relative to a 30-year old female with no risk factors 2 to 5 times and do not vary substantially with age. Nonetheless, the DeCaprio et al. model implies that the increase in relative vulnerability to severe illness will result, in large part, from the number of conditions one has at any age.

Three final points about Figure 1. First, age, *per se*, has a very small impact on relative vulnerability. That is, growing older, when one has no risk factors, has virtually no impact on vulnerability. It is the accumulation of risk factors and the interactions of these risk factors with age that lead to higher vulnerability at older ages. Second, a male, who has no other health-related risk factors, is at slightly higher vulnerability to severe illness compared to females and this male-female differential in vulnerability rises modestly with age, so that, by age 72, a male’s vulnerability, relative to a 30-year old female, is 1.14 times higher. Third, the distribution of and disparities in the relative vulnerability to severe illness from COVID-19 in the U.S. adult population reported below will be driven by the marginal contributions of individual risk factors displayed in Figure 1 based on the DeCaprio et al. model and by the prevalence of these risk factors in the population.

## RESULTS

### Prevalence of Health-Related Risk Factors in the United States and their Disparities

In Table 2, we present the prevalence of the risk factors and how they vary by age, demographic group and socioeconomic status. We test whether differences within each group are statistically significant from the base category (age 25-44, NH White, BA or more, top income quartile) and denote these with asterisks.

**Table 2.** Age, Race-Ethnicity, and SES in Prevalence (%) of Health-Related Risk Factors

| Risk factor | Overall | Age |  |  | Race-ethnicity |  |  | Education |  | Household Income |  |
| --- | --- | --- | --- | --- | --- | --- | --- | --- | --- | --- | --- |
|  |  | 25-44 | 45-64 | 65+ | NH White | NH Black | Hispanic | HS or less | BA or more | Bottom Quartile | Top Quartile |
| Hypertension | 33 | 14 | 35*** | 59*** | 33 | 44*** | 23*** | 39*** | 26 | 46*** | 25 |
| Diabetes | 12 | 4 | 14*** | 23*** | 11 | 16*** | 14* | 15*** | 9 | 18*** | 8 |
| Asthma | 11 | 12 | 10** | 9*** | 11 | 13** | 9** | 11* | 10 | 15*** | 9 |
| Hospitalization | 10 | 7 | 9** | 17*** | 10 | 12* | 9 | 12*** | 7 | 18*** | 6 |
| Cancer | 9 | 2 | 7*** | 21*** | 10 | 6*** | 5*** | 9 | 9 | 11** | 8 |
| Lung disease | 6 | 3 | 6*** | 11*** | 6 | 7 | 3*** | 9*** | 3 | 12*** | 2 |
| Heart disease | 6 | 1 | 5*** | 14*** | 6 | 7 | 2*** | 7*** | 4 | 11*** | 3 |
| Severe obesity | 5 | 5 | 6 | 4* | 4 | 9*** | 5 | 6*** | 3 | 7*** | 3 |
| Heart attack | 4 | 1 | 4*** | 10*** | 5 | 5 | 2*** | 6*** | 2 | 7*** | 2 |
| Stroke | 3 | 1 | 3*** | 8*** | 3 | 6*** | 2* | 4*** | 2 | 7*** | 2 |
| Neurological conditions | 3 | 1 | 2*** | 5*** | 3 | 2** | 2*** | 4*** | 1 | 6*** | 0 |
| Chronic Kidney Disorder | 1 | <1 | 1** | 1*** | 1 | 1 | <1* | 1 | 1 | 1 | 0 |
| Sample Sizes | 13,150 | 6,808 | 4,392 | 1,950 | 7,082 | 4,158 | 1,464 | 5,262 | 4,406 | 2,550 | 3,774 |

Data Source: 2017 Wave, PSID. Sample: PSID heads and spouses, 25 years and older. Weights: PSID individual cross-section weights.
Income quartiles are determined across households using family weights.
**Notes:** Asterisks (\*) are for tests of risk factors of a subgroup being significantly different from base group, where: \* P-value $\leq 0.10$ ; \*\* P-value $\leq 0.05$ ; \*\*\* P-value $\leq 0.01$ . Base groups are 25-44, NH White, BA or more, top income quartile.

The risk factors are listed in order of their prevalence in the overall U.S. adult population based on the PSID data. Hypertension is the most prevalent factor, at 33%. Hypertension increases with age, with 59% of those 65 and older having been diagnosed. Diabetes and asthma are the second and third most prevalent risk factors at 12% and 11%, respectively. Diabetes increases with age, with only 4% of those in early adulthood (25-44 years old) having been diagnosed while one quarter (23%) of those 65 and older have the disease. In contrast, asthma declines with age, going from 12% for those 25-44 years old to 9% for those 65 and older. Some 9% of the adult population has been diagnosed with cancer and its prevalence rises with age up to 1 in 5 (21%) of those 65 and older. Five percent of adults are severely obese and there is little variation by age. Among the remaining health-related risk factors, the prevalence ranges from 1% to 6%, all increasing with age. Finally, 10% of adults report being hospitalized in the preceding year for one night or more, including 17% of those 65 and older.

As the other columns in Table 2 make clear, there is an unequal distribution of these health-related risk factors by race-ethnicity, educational attainment, and income. Compared to NH Whites, NH Blacks have higher prevalence of most of the risk factors for COVID-19, with nearly all of these differences being statistically significant. For example, NH Blacks are one third more likely to have been diagnosed with hypertension than NH Whites (44% vs 33%). Cancer is more prevalent for NH Whites than NH Blacks (10% for NH Whites vs 6% for NH Blacks). Neurological conditions are also more prevalent for NH Whites (3%) than NH Blacks (2%). Prevalence rates for the risk factors listed in Table 2 are the same or lower for Hispanic adults than NH Whites (or NH Blacks), except for diabetes, which is more prevalent for Hispanics than NH Whites. The prevalence of hypertension is 23% for Hispanics compared to 33% for NH Whites. These race-ethnic differences have been documented in prior studies (32–34).

The prevalence of risk factors is also quite different by education. Adults in the United States with no more than a high school education have higher rates for almost all risk factors compared to adults with a college degree. For example, 39% suffer from hypertension compared to one quarter (26%) of the higher educated group. Two notable exceptions to this pattern are cancer and chronic kidney disease, which do not vary by education.

Adults in the bottom quartile of household income have higher rates for every risk factor compared to those in the top quartile, and these differences are sizeable. Compared to those in the top income quartile, those in the bottom quartile are about twice as likely to have been diagnosed with hypertension (25% vs 46%), diabetes (8% vs 18%) and asthma (9% vs 15%), are about three times more likely to have had a heart attack (2% vs 7%) or a stroke (2% vs 7%) and are six times more likely to suffer from lung disease (2% vs 12%). Those in the lowest income group are three times more likely to have had an in-patient hospitalization in the past year (6% vs 18%).

Table 2 also shows that the prevalence of nearly all risk factors increases with age. The total number of risk factors also increases with age. In Figure 2, we display the percent of the population with 0, 1, 2 and 3 or more of the risk factors listed in Table 2 across three age groups. Among all adults, 47% have 0 risk factors, while 13% have 2 and another 13% have 3 or more, for an average of 1.02 risk factors in the U.S. adult population. The number of risk factors increases across age groups, with 64% of adults 25-44 having no risk factors while only 20% of those 65 and older having none. Only 3% of younger adults have 3 or more risk factors, while 28% of those 65 and older have 3 or more.

**Figure 2.**
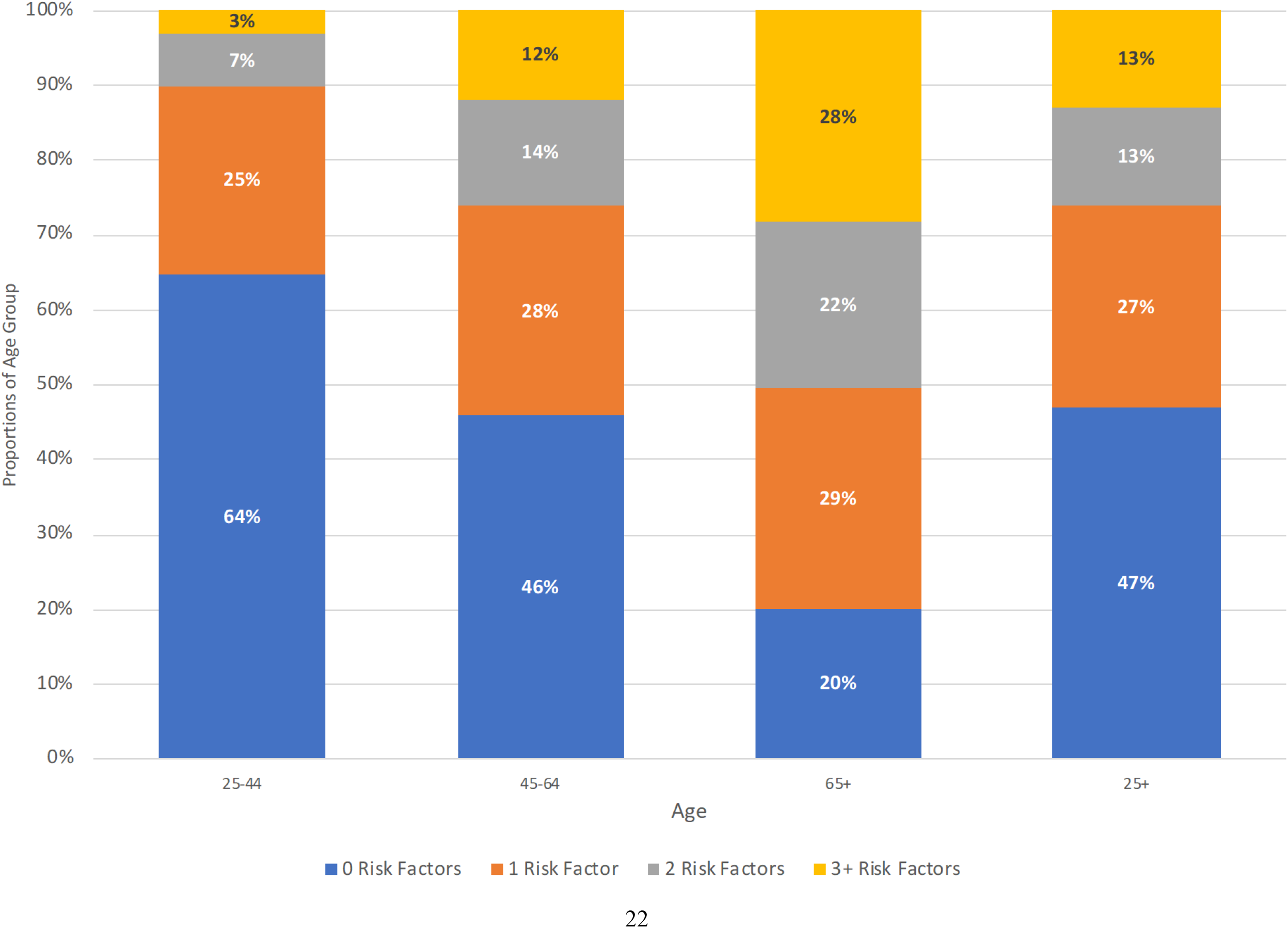
Distribution of Number of Risk Factors by Age

We showed that the distribution of age varies across race-ethnicity, education, and income groups in our sample. Appendix Tables C-1 - C-3 report the prevalence of each of the risk factors by race-ethnicity, socioeconomic status, and age. Differences in the prevalence of hypertension between NH Blacks and NH Whites and by education and income emerge early in life and are large by midlife. Hispanics have a health advantage in hypertension in midlife but the prevalence of diabetes among Hispanics is greater than for NH Whites at ages 45-64. There are disparities across nearly all conditions for less-educated and low-income Americans that are similar to those for NH Blacks. These disparities become large in midlife though there are statistically significant differences by education and income in the prevalence of nearly all health conditions among those age 25-44.

In Figure 3, we present the distribution of the number of risk factors for the overall population and by age across race-ethnic groups, educational attainment, and household income, using the same format as Figure 2. Consistent with the disparities by race-ethnicity in individual risk factors, among all adults, NH Blacks are more likely to have three or more risk factors compared to NH Whites (18% vs 13%), while only 7% of Hispanics have such a high number (Figure 3(a), 25+ age group). Among adults with 12 years of schooling or less, 17% have 3 or more risk factors, compared to just 7% of those with a college degree (Figure 3(b), 25+ age group). Finally, the differences in numbers of risk factors are particularly dramatic by income, with 24% of adults in the bottom quartile of household income having 3 or more risk factors compared to only 6% among those in the top income quartile (Figure 3(c), 25+ age group). All of the differences highlighted above are statistically significant.

**Figure 3.**
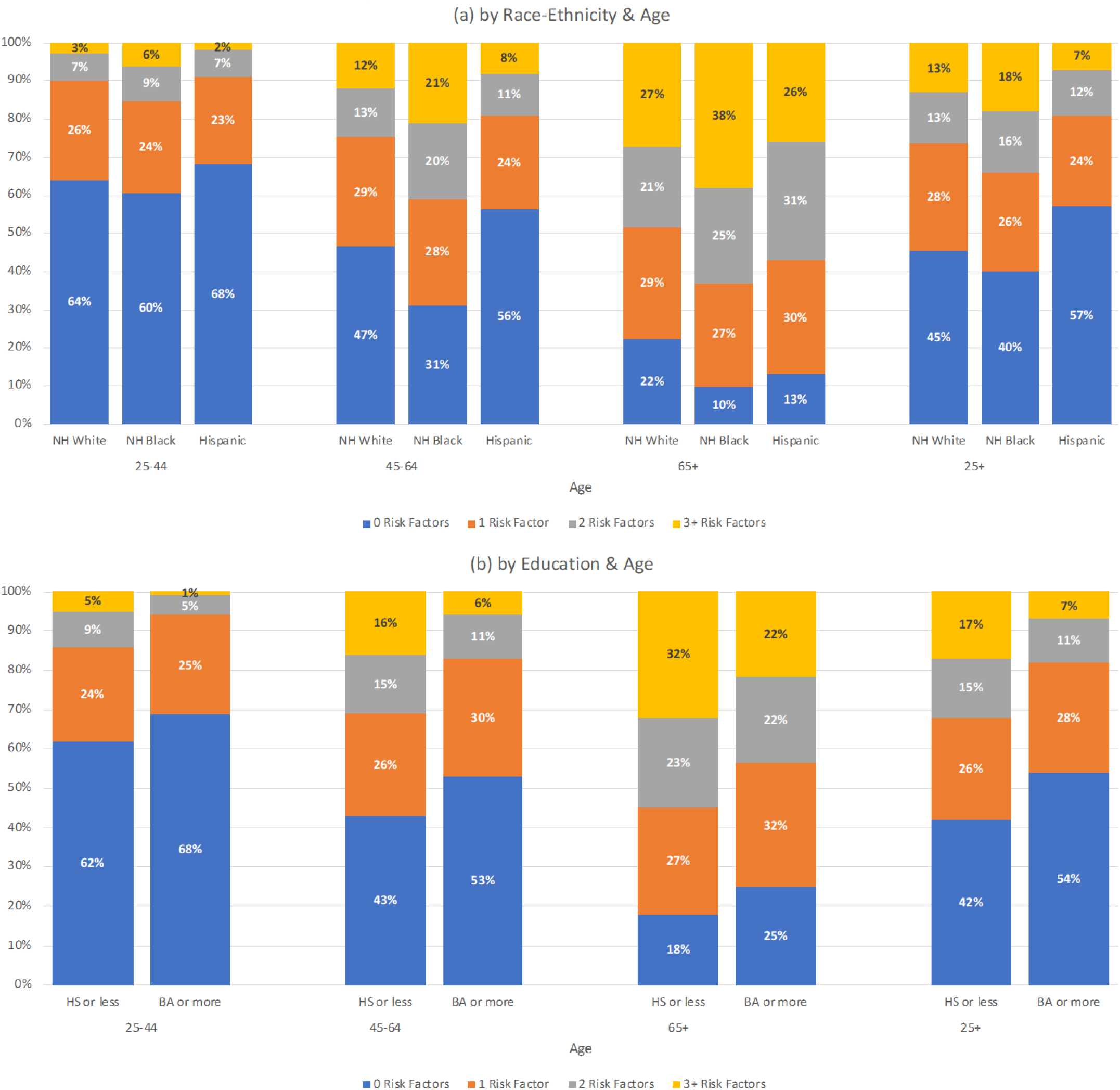
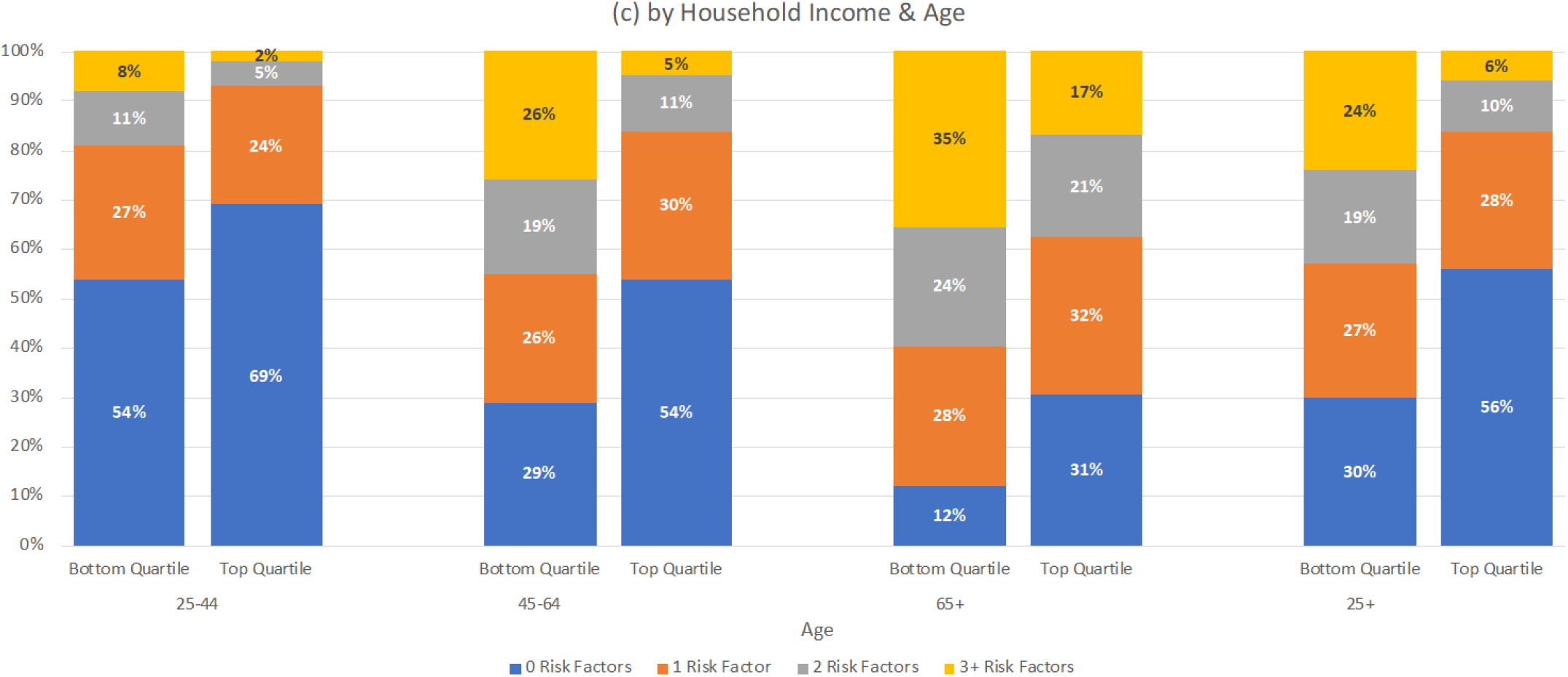
Distribution of Number of Risk Factors

Figure 3 also shows the distribution of the number of risk factors by age across race-ethnicity, education, and household income. For NH Blacks, those with 12 years of schooling or less, and those in the bottom quartile of income, risk factors accumulate at earlier ages. For example, by age 45-64, 41% of NH Blacks have two or more risk factors compared to only 25% of NH Whites. On average, fewer adults age 45-64 with household incomes in the top income quartile have two or more conditions (16%) than those age 25-44 in the bottom income quartile (19%), and fewer adults in the oldest group with household incomes in the top quartile have two or more conditions (38%) than those in middle age (45-64) with incomes in the bottom quartile (45%). All of these differences highlight above are statistically significant.

### Disparities in Relative Vulnerability to Severe Illness in the United States

In Figure 4, we display median values of the relative vulnerability index by numbers of risk factors for the three age groups and the overall adult population.^6^ Recall that the index measures the odds of having severe illness from COVID-19 relative to a 30 year-old female with no risk factors. The median value is 4.08 (not shown in Figure 4). As seen in Figure 4, relative vulnerability varies substantially by number of risk factors and by age. Among all ages (the 25+ group), those with no risk factors are 1.9 times more vulnerable compared to a 30 year-old female. As the number of risk factors increases, relative vulnerability increases, with the median risk of individuals with 3 or more risk factors at 47.9 times greater odds of becoming severely ill. On average, individuals in this high-risk group have 3.8 risk factors and 9% have 5 or more risk factors. Individuals with particular risk factors are also likely to have more risk factors. For example, adults with hypertension had, on average, 1.2 other risk factors, while those who did not have hypertension had an average of 0.4 risk factors.

**Figure 4.**
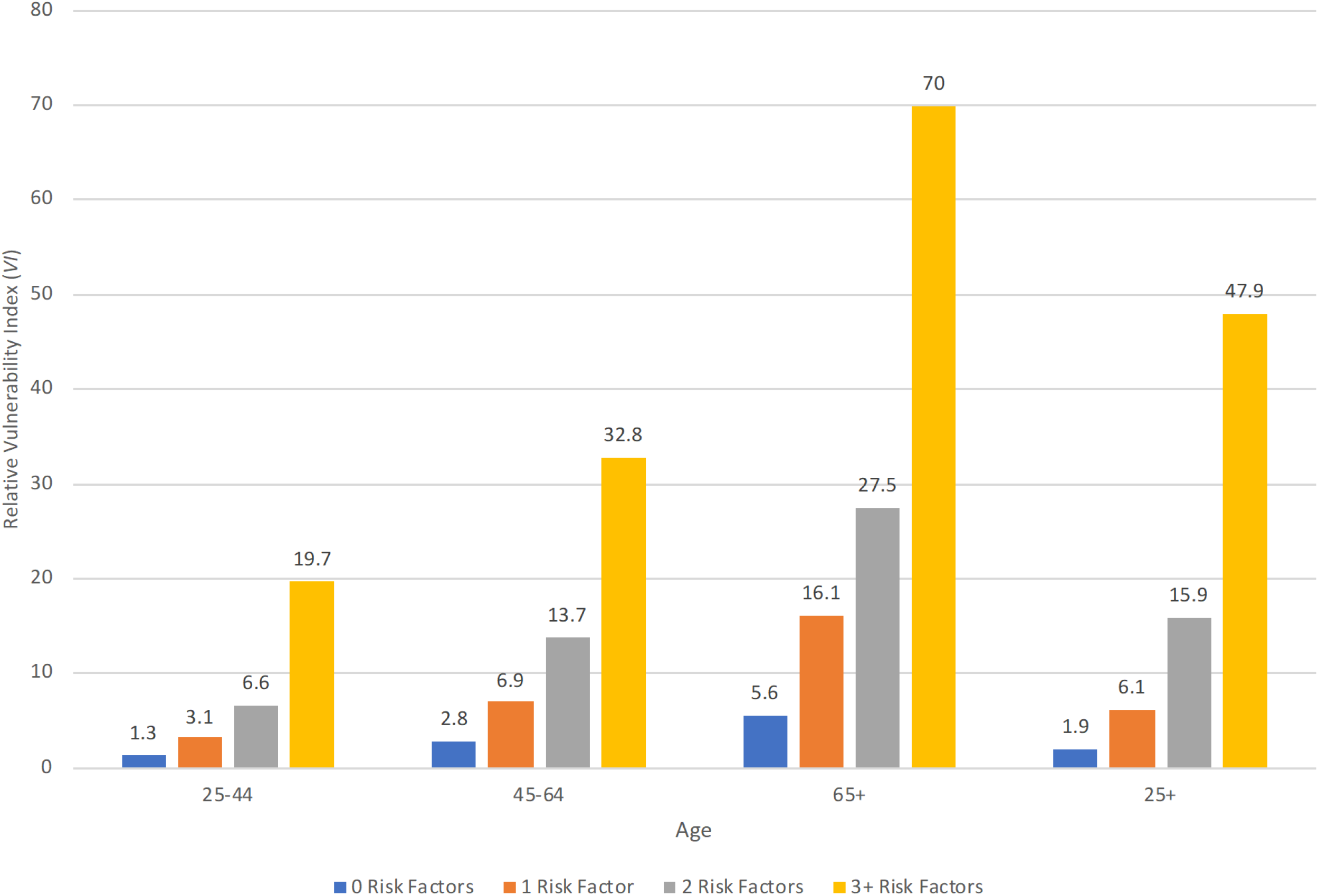
Median Relative Vulnerability Index (*VI*) by Number of Risk Factors & Age

Figure 4 also makes clear that the increasing numbers of risk factors for those at older ages displayed in Figure 2, result in a progressive increase in relative vulnerability across age groups.

For example, individuals who are 65 and older and who have 3 or more risk factors are 70 times more likely to become severely ill than a 30 year-old healthy (no risk factors) female. Among individuals in this high-risk group, 25% have 5 or more risk factors and half are age 75 or older. While this estimate of relative vulnerability is extraordinarily high, the relative magnitudes of the differences by age and number of risk factors illustrate how the accumulation of risk factors with age makes many older Americans much more vulnerable to COVID-19.

Figure 4 also illustrates how having multiple risk factors heightens the vulnerability to severe illness from COVID-19 at any age. For example, adults 65 and older with no risk factors (20% of that age group) have a median vulnerability that is 5.6 times that of the reference person (30 year-old female with no risk factors). This is almost the same relative vulnerability to severe illness as for 25-44 year-olds with 2 risk factors (6.6 relative vulnerability and 7% of the younger age group). In fact, the relative vulnerability of those 65 and older with no risk factors is more than 3 times lower than the relative vulnerability of 25-44 year-olds with 3 or more risk factors. This comparison illustrates both the disturbing consequences of younger adults having multiple risk factors and the apparent benefits to older age adults of having fewer risk factors with respect to vulnerability of becoming severely ill from COVID-19.

The results in Figure 4 for relative vulnerability represent the differences in the distributions of numbers of risk factors for different age groups *and* the impact that particular risk factors have on relative vulnerability at different ages. Recall for example that Figure 1 shows that hypertension and lung disease have the largest impacts on the relative vulnerability scores at all ages and that the impacts of both increased markedly with age. At the same time, as we noted in our discussion of Table 2, the prevalence of hypertension in the U.S. population is high (33%) and increases across age groups, while, relatively speaking, the prevalence of lung disease is much lower (6%) and increases less markedly with age. Thus, the relative vulnerability index allows one to characterize the combined impact of (and differences in) the prevalence of risk factors and their impact on population-level vulnerability, providing greater insight than simply looking at the prevalence of risk factors can provide.

In Figure 5 we display the median relative vulnerability scores by age for race-ethnic, education, and income groups. Recall that in Figure 3(a) we found that NH Blacks have more risk factors, on average, than NH Whites, and that NH Blacks have a higher number of risk factors after age 45 than NH Whites. The higher prevalence of hypertension among NH Blacks contributes to this race differences in the number of risk factors (see Appendix Table C-1). We also found that the prevalence of most risk factors is lower among Hispanics, other than diabetes (see Appendix Table C-1). But diabetes has a smaller effect on VI than hypertension (Figure 1). As shown in Figure 5(a), by age 45, sizeable differences in relative vulnerability to severe illness emerge between NH Blacks and NH Whites and these gaps remain large at older ages. Hispanics have lower relative vulnerability at all ages. Across the whole sample (25+), the difference in relative vulnerability between NH Blacks and NH Whites is muted. This is due to the age distributions shown in Table 1 where those who are age 65 and older make up 28% of the population of NH Whites but only 17% of the population of NH Blacks.

**Figure 5.**
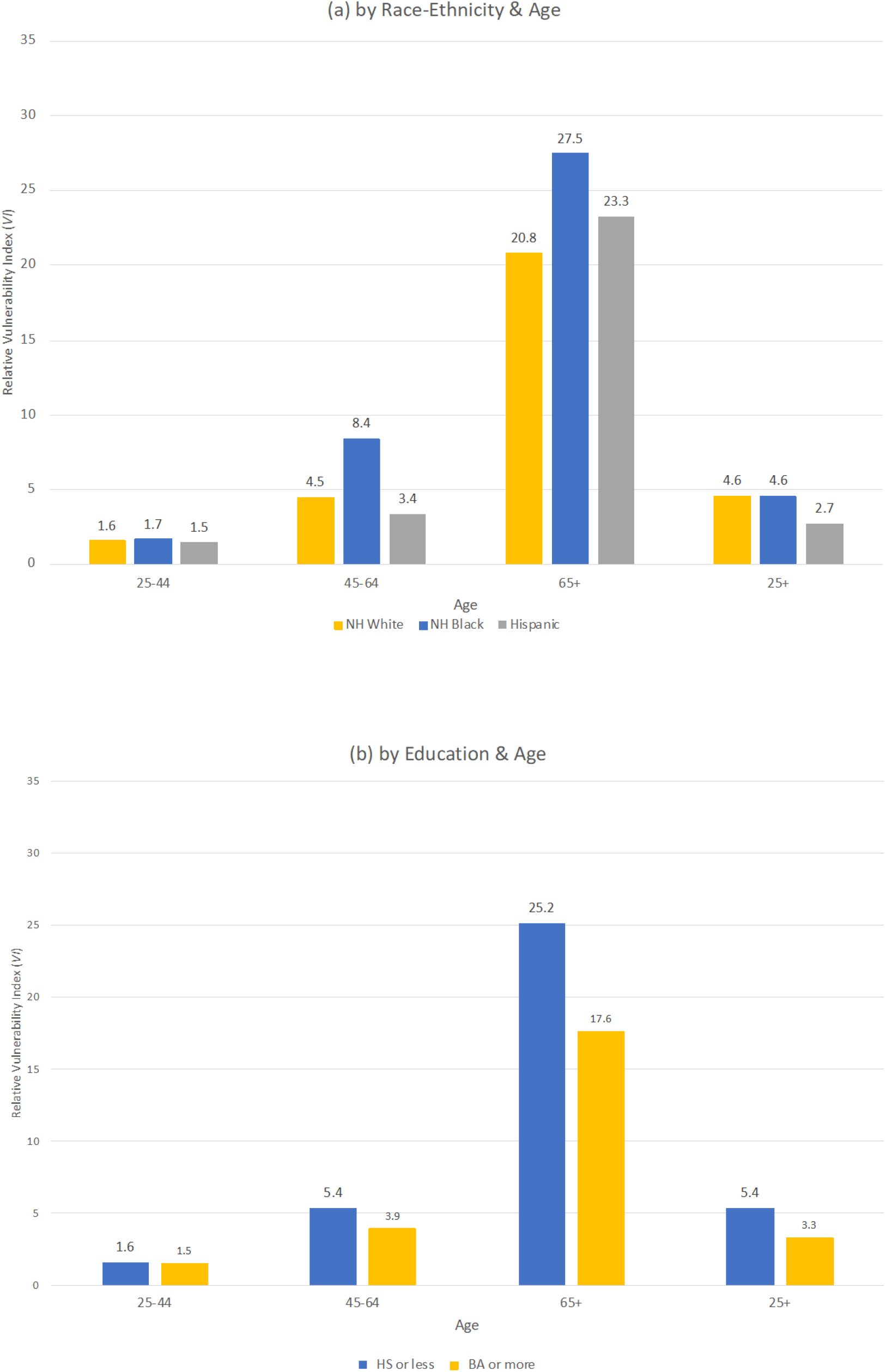
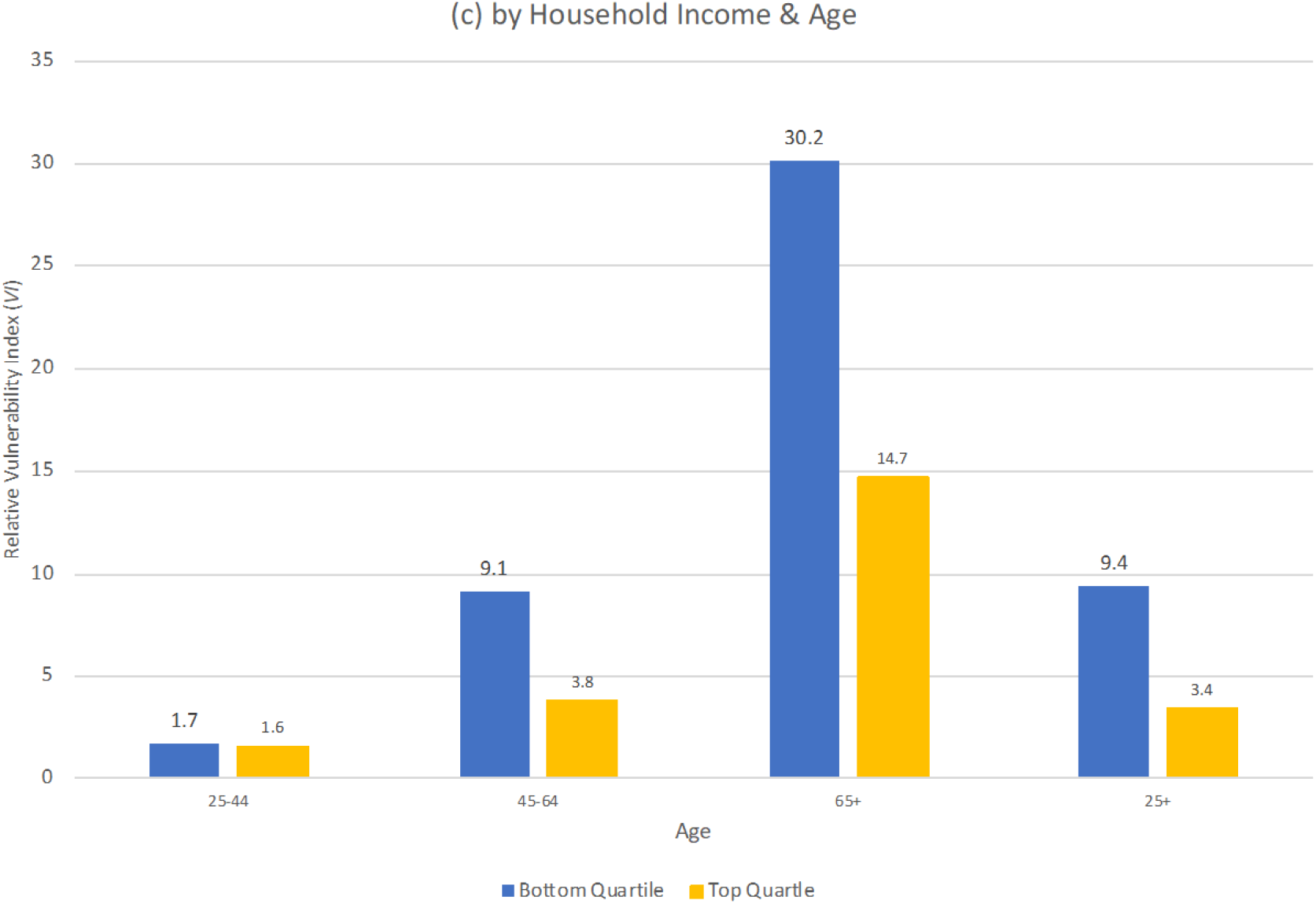
Median Relative Vulnerability Index (*VI*)

Figure 5(b) has the same format for showing the median relative vulnerability among adults with a high school degree or less education and those who are college educated. On average, relative vulnerability is 60% higher for those with high school or less (5.4) than for those with a college degree (3.3), and this gap by education emerges in midlife.

Finally, Figure 5(c) presents the median vulnerability for those in the bottom and top quartiles of household income. Among all adults (ages 25+), there is nearly a 3-fold difference in relative vulnerability, with those in the bottom quartile of income having 9.4 higher odds of severe illness compared to the reference person, while for those in the top income quartile, the odds are only 3.4 times higher. This gap by income in relative vulnerability emerges among middle-age adults and reflects large differences in prevalence of risk factors (Figure 3(c)) by income.

A full assessment of the relationships among race-ethnicity, education, and income is beyond the scope of this paper. But in light of the correlations among these dimensions of population subgroups, we examined differences in vulnerability in midlife across race-ethnic groups within education and income groups, the differences in vulnerability across education within race-ethnic and income groups, and the differences in vulnerability across income within race-ethnic and education groups (see Appendix Table C-4). Taking account of socioeconomic differences mutes, but does not eliminate the greater vulnerability of NH Blacks compared to NH Whites. Hispanics’ health advantage in vulnerability is pronounced among low-income adults. Education and income differences in vulnerability remain large but are reduced within race-ethnic groups.

## DISCUSSION

The DeCaprio et al. model predicts hospitalizations due to respiratory infections prior to the COVID-19 outbreak and as a function of age and preexisting health risk factors. How accurately does this model predict actual disparities by age, race-ethnicity, and socioeconomic status in hospitalizations associated with the coronavirus? We address this question using early surveillance data from the CDC.

Hospitalization is influenced by two channels: exposure and susceptibility to infection and its complications. Each may differ in their determinants and, thus, may differ across particular viruses and diseases. Susceptibility and complications are likely to depend heavily on the sort of preexisting conditions included in the DeCaprio et al. model. Evidence from New York City indicates that, conditional on testing positive for COVID-19, the comorbidities and the demographic characteristics of patients hospitalized with COVID-19 are similar to those admitted with other acute respiratory infections (11). However, exposure to COVID-19 likely differs by factors other than age and preexisting health conditions. Separating these channels empirically is beyond the scope of available data. However, we can compare the model’s predictions for disparities in vulnerability presented above with the distributions of available characteristics of those that have been hospitalized to date with severe complications from the coronavirus.

Garg et al. (20) report on the prevalence of preexisting health conditions disaggregated by age for laboratory-confirmed COVID-19 hospitalizations during March 2020. We predict *VI* based on the average conditions of these hospitalized patients overall and by age group. We compare *VI* based on the average preexisting conditions of hospitalized patients to the distribution of *VI* in the PSID. Appendix D describes these calculations in more detail.

The *VI* of hospitalized patients of all ages is 8.51, which corresponds to the 66% percentile of *VI* for the PSID sample. For hospitalized patients between 18-49 it is 3.04, for those age 50-64 it is 7.87, and for those 65 and older it is 23.14, corresponding to the 48^th^ percentile, the 65^th^ percentile, and the 84^th^ percentile of the PSID vulnerability distribution, respectively. This correspondence between the *VI* for actual COVID-19 hospitalizations and that in the PSID based on the DeCaprio et al. model is consistent with our findings that older adults with multiple preexisting conditions in the general population are more vulnerable to severe complications requiring hospitalization and that a substantial fraction of adults in midlife are also vulnerable to hospitalization. For younger adults, our *VI* estimates based on health conditions correspond less well to actual COVID-19 hospitalizations. This lack of correspondence suggests that, compared to older adults, exposure to infection is relatively more important for younger adults than susceptibility to infection and complications.

The distinction between exposure and infection also sheds light on race-ethnic disparities in observed hospitalizations. The CDC data indicate a dramatic over-representation of NH Blacks among hospitalized patients while we find no NH Black-NH White difference in median *VI* when all ages are combined (see Figure 5a) (21). Within age groups, we report that NH Blacks have higher *VI*. These patterns are consistent with higher rates of exposure among NH Blacks than NH Whites that are compounded by disparities in health conditions within age groups. Relative to NH Whites, Hispanics are also overrepresented among patients hospitalized with COVID-19, both overall and after adjusting for age (21). We report that Hispanics have lower median *VI* than NH Whites both overall and within age groups, and thus rates of exposure are likely higher for Hispanics than NH Whites.

## CONCLUSION

This paper provides the first nationally representative estimates of vulnerability to severe complications from COVID-19 overall and across race-ethnicity and socioeconomic status. We use the PSID to examine the prevalence of specific health conditions associated with complications from COVID-19 and to calculate, for each individual, an index of the risk of severe complications from respiratory infections developed by DeCaprio et al. (23) based on these health conditions. We show large disparities across race-ethnicity and socioeconomic status in the prevalence of conditions associated with adverse outcomes, and in the overall risk of severe complications. Moreover, we show that these disparities emerge early in life, prior to age 65.

Consistent with the surveillance data, our results suggest a higher risk of adverse outcomes in midlife for non-Hispanic Blacks. Education and income are not included in the surveillance data but our results further suggest that adults with a high school degree or less and low-income Americans are also at high risk of adverse outcomes starting in midlife. These results are especially important as states and municipalities start to reopen businesses and public services as recent evidence suggests that disadvantaged groups are less likely to be able to socially distance at work (27). The evidence that we present shows large disparities in preexisting health conditions among working-age adults for these same groups. Combined, these results suggest that localities with high levels of poverty, a high percentage of African-Americans, or a high concentration of less-educated adults face potentially devastating effects of the virus.

There are caveats to this study. First, we are likely understating the disparities in the risk for severe complications from COVID-19 by race-ethnicity, education, and income based on preexisting health conditions because disadvantaged populations have higher rates of undiagnosed diseases (35–39), because socioeconomic status and race-ethnicity affect whether an individual is hospitalized regardless of preexisting conditions (40), and because among those diagnosed with a disease, rates of control for some diseases are lower for disadvantaged populations (36, 37, 39, 41).

Second, there is more work to be done exploring interactions of race-ethnicity, education, and income. There is ample evidence of large mortality gaps across these groups (42), and this is an area for more extensive exploration.

Finally, we have discussed why disparities in the risk of severe complications based on preexisting health conditions does not capture the full risk of hospitalization and death from COVID-19 because it does not account for factors influencing exposure to the virus or access to high quality care. All of these factors, including the preexisting health conditions we examine, are influenced by systemic inequalities in society and in our health care system. Such inequalities will make it difficult to isolate the influence of health conditions on disparities in the overall effect of COVID-19 pandemic on the U.S. population but starting to disentangle these effects is crucially important.

## Data Availability

All data used in this study is publicly available at https://simba.isr.umich.edu/.

## Funding Acknowledgement

This paper was prepared with support, in part, from the Aging Studies Institute at Syracuse University, the Duke Center for Population Health and Aging, which receives core support (P30AG034424) from the National Institute on Aging, and by the California Center for Population Research at the University of California at Los Angeles, which receives core support (P2C-HD041022) from the Eunice Kennedy Shriver National Institute of Child Health and Human Development. The collection of data used in this study was partly supported by the National Institutes of Health under grant number R01 HD069609 and R01 AG040213, and the National Science Foundation under award numbers SES 1157698 and 1623684.

## Appendix A Comparison of PSID Sample with Current Population Survey (CPS)

**Table A-1.** Percent of Demographic and SES Groups by Age across Age Categories & Mean Age for PSID Sample

|  | Percent across Age Categories |  |  | Percent within Age Categories |  |  | Prop (%),<br>All Ages<br>(25+) | Mean<br>Age | Sample<br>sizes |
| --- | --- | --- | --- | --- | --- | --- | --- | --- | --- |
|  | 25-44 | 45-64 | 65+ | 25-44 | 45-64 | 65+ |  |  |  |
| Overall |  |  |  | 37.6 | 38.1 | 24.3 |  | 51.9 | 13,150 |
| Gender |  |  |  |  |  |  |  |  |  |
| Female | 37 | 38 | 26 | 51 | 52 | 55 | 52 | 52.4 | 7,140 |
| Male | 39 | 38 | 23 | 49 | 48 | 45 | 48 | 51.5 | 6,010 |
| Race and Ethncity |  |  |  |  |  |  |  |  |  |
| NH White | 33 | 38 | 28 | 64 | 72 | 83 | 71 | 53.6 | 7,082 |
| NH Black | 42 | 41 | 17 | 12 | 12 | 7 | 11 | 49.2 | 4,158 |
| Hispanic | 52 | 36 | 13 | 18 | 12 | 7 | 13 | 46.5 | 1,464 |
| NH Other | 52 | 32 | 15 | 7 | 4 | 3 | 5 | 47.4 | 446 |
| Education |  |  |  |  |  |  |  |  |  |
| HS or less | 32 | 40 | 28 | 32 | 41 | 44 | 38 | 54.3 | 5,262 |
| Some college | 39 | 38 | 23 | 25 | 24 | 22 | 24 | 51.2 | 3,482 |
| BA or more | 43 | 36 | 22 | 43 | 36 | 34 | 38 | 50.1 | 4,406 |
| Sample sizes | 6,808 | 4,392 | 1,950 |  |  |  |  |  |  |

**Data Source:** PSID, 2017 Wave. **Sample:** Household heads and spouses, ages 25 and older. **Weights:** PSID individual cross-section weights.

**Table A-2.** Percent of Demographic and SES Groups by Age across Age Categories & Mean Age for CPS Sample

|  | Percent across Age Categories |  |  | Percent within Age Categories |  |  | Prop (%),<br>All Ages<br>(25+) | Mean<br>Age | Sample<br>sizes |
| --- | --- | --- | --- | --- | --- | --- | --- | --- | --- |
|  | 25-44 | 45-64 | 65+ | 25-44 | 45-64 | 65+ |  |  |  |
| Overall |  |  |  | 34.8 | 40.3 | 24.9 |  | 52.4 | 101,462 |
| Gender |  |  |  |  |  |  |  |  |  |
| Female | 35 | 40 | 25 | 53 | 52 | 54 | 53 | 52.5 | 53,840 |
| Male | 35 | 41 | 25 | 47 | 48 | 46 | 47 | 52.4 | 47,622 |
| Race and Ethncity |  |  |  |  |  |  |  |  |  |
| NH White | 30 | 41 | 29 | 60 | 70 | 80 | 69 | 54.3 | 65,151 |
| NH Black | 38 | 43 | 20 | 12 | 11 | 9 | 11 | 50.6 | 11,378 |
| Hispanic | 49 | 38 | 13 | 19 | 12 | 7 | 13 | 46.8 | 15,976 |
| NH Other | 47 | 37 | 16 | 10 | 7 | 5 | 8 | 48.1 | 8,957 |
| Education |  |  |  |  |  |  |  |  |  |
| HS or less | 29 | 41 | 30 | 30 | 38 | 44 | 37 | 55.0 | 38,019 |
| Some college | 35 | 41 | 23 | 28 | 28 | 25 | 27 | 51.8 | 27,560 |
| BA or more | 40 | 39 | 21 | 42 | 35 | 31 | 36 | 50.4 | 35,883 |
| Sample sizes | 38,732 | 40,401 | 22,329 |  |  |  |  |  |  |

**Data Source:** CPS, 2017 Annual Social & Economic Supplement. **Sample:** Household heads and spouses, ages 25 and older. **Weights:** ASEC weights.

## Appendix B Formula for the DeCaprio et al. Model and Corresponding Risk Factors in the PSID

In Table B-1, we provide the variable names and question wording from the 2017 Wave of the PSID for these risk factors. Table B-2 displays the variables used and coefficient estimates for the DeCaprio et al. Model^7^ along with the corresponding health-related risk factors that are available in the PSID.

PSID staff has compared self-reports about health conditions to the gold-standard for self-reported health conditions, the National Health Interview Survey (NHIS), to evaluate the quality of the PSID data. The PSID estimates for nearly all measures are close to those in the NHIS, a personal household interview study that has been used by the National Center for Health Statistics to monitor Americans’ health since 1957. PSID estimates also show time trends consistent with those from the NHIS (43). There are two exceptions. Chronic kidney disease is twice as common in the NHIS than in the PSID, in which 1% report kidney disease (44). The PSID information about kidney disease, unlike all other conditions we examine, comes from reports about “other serious chronic conditions,” without a specific prompt for kidney-related conditions. The NHIS information is obtained from a specific question about “weak or failing kidneys,” and specific questions are more likely to elicit these reports. The other exception is the prevalence of “neurocognitive conditions,” which we indicate by whether sample members were ever told by a doctor or health professional that they had “permanent loss of memory or mental ability.” However, the PSID self-reported measure has external validity as demonstrated by its consistent positive association with the Eight Item Interview to Differentiate Aging and Dementia Screen (AD8) (45). Though Insolera & Freedman (43) validate estimates of obesity (BMI ≥ 30) with NHIS, the estimates of severe obesity (BMI ≥ 40) in Table 2 match those in the BRFSS. Rates of hypertension by race-ethnicity in the PSID match crude rates from NHANES almost exactly (34).

As noted in the text, the PSID does not have all of the variables that DeCaprio et al.(23) used in the estimation of the logistic regression for this Survey Model. In our implementation of the model, we assumed that the indicators for the risk factors not collected in the PSID had a value of 0. As we note in the text, this means that our estimates of the vulnerability index scores of individuals in the PSID understate their risk of serious disease. The extent of this understatement and how the extent of understatement will vary in the population depends on both the prevalence of the particular risk factors in the population and their marginal impacts on the vulnerability index score, which is determined by the coefficients on these variables.

The following conditions are not included: sickle cell disease (SCD), hemodialysis, pneumonia, rheumatic heart disease, and liver disease (cirrhosis of the liver). The prevalence and/or incidence of most of these conditions is low. SCD affects approximately 100,000 Americans. It is more common among Blacks, than other race-ethnic groups, but occurs among under 0.3% of Black births. (46). In 2017, 746,557 Americans had kidney failure and needed dialysis or a kidney transplant to survive (47). About 250,000 people in the United States seek care each year in a hospital due to pneumonia (48). Finally, the incidence of rheumatic fever is less than 2 cases per 100,000 school-age children and most cases occur in children 5 – 15 years of age (49). The prevalence of liver disease is higher. About 4.5 million adults are diagnosed with liver disease, with a prevalence of 1.8% (50). Thus, with the exception liver disease (cirrhosis of the liver), the risk factors in the DeCaprio et al. model, but not in the PSID, are of very low prevalence and/or incidence. Liver disease has a very low coefficient estimate and a small interaction effect with age. Sickle cell anemia, which has a fairly sizeable impact in the calculation of the index for NH Blacks at ages up to around 50 has a low prevalence in the population. However, because the prevalence of Sickle cell anemia cannot be determined for our PSID sample, the values for the vulnerability index in the PSID data will be (slightly) understated, on average, for NH Blacks and some Hispanics.

**Table B-1.** Health-Related Risk Factors and Other Variables, Variable Names, and Question Wording from the 2017 Wave of the PSID

| **Risk Factor/Variable** | **PSID Variable** | **Question Wording** |
| --- | --- | --- |
| Age | ER34504 | What is your current age? |
| Gender | ER32000 | What is your gender? |
| Past Hospitalization | ER68511, ER69638 | Were you a patient in a hospital overnight or longer at any time during 2016? |
| Lung Disease | ER68454, ER69581 | Has a doctor or other health professional ever told you that you had chronic lung disease such as bronchitis or emphysema? |
| Asthma | ER68449, ER69576 | Has a doctor or other health professional ever told you that you had asthma? |
| Severe Obesity | ER68568, ER68569, ER69695, ER69696, ER68566, ER68567 | Constructed BMI using height and weight variables and designating severely obese if BMI ≥ 40 |
| Diabetes | ER68459, ER69586 | Has a doctor or other health professional ever told you that you had diabetes or high blood sugar? |
| Hypertension | ER68444, ER69571 | Has a doctor or other health professional ever told you that you had high blood pressure or hypertension? |
| Heart Disease | ER68439, ER69566 | Has a doctor or other health professional ever told you that you had coronary heart disease, angina, congestive heart failure? |
| Heart Attack | ER68433, ER69560 | Has a doctor or other health professional ever told you that you had a heart attack? |
| Stroke | ER68427, ER69554 | Has a doctor or other health professional ever told you that you had a stroke? |
| Kidney Disease | ER68498, ER69625 | Is there any other serious, chronic condition that a doctor or other health professional ever told you had? [ 3: Kidney disease] |
| Cancer | ER68479, ER69606 | Has a doctor or other health professional ever told you that you had cancer or a malignant tumor? |
| Neurological condition | ER68469, ER69596 | Has a doctor or other health professional ever told you that you had permanent loss of memory or mental ability? |

**Table B-2.** Risk Factors & Other Conditions from the DeCaprio et al. Model

| **Variables in DeCaprio et al. Survey Model** | **Coefficient Estimate** | **Variables available in the PSID** |
| --- | --- | --- |
| intercept | -6.740 |  |
| Age | 0.041 | Age |
| Male | 0.171 | Male |
| Prior Admissions | 0.682 | Hospitalized |
| Prior ER Visits | 0.413 |  |
| Chronic obstructive pulmonary disease (COPD) or emphysema, cystic fibrosis, or chronic bronchitis | 1.167 | Lung Disease |
| Asthma | 1.393 | Asthma |
| Obesity (BMI $\geq 40$) | 0.935 | Severe Obesity (BMI $\geq 40$) |
| Diabetes (other than when you were pregnant) | 0.096 | Diabetes |
| Hypertension (or high blood pressure) | 0.832 | Hypertension |
| Congestive Heart Failure | 0.982 | Heart Disease |
| Heart attack | 0.159 | Heart Attack |
| Rheumatic heart disease | 0.788 |  |
| Stroke | 0.285 | Stroke |
| Sickle cell anemia/HIV infection/Transplant | 2.582 |  |
| Chronic kidney disease | 0.966 | Kidney Disease |
| Hemodialysis | 1.369 |  |
| Liver disease | 0.055 |  |
| Pneumonia, acute bronchitis, influenza or other acute respiratory infection | 0.696 |  |
| Cancer | 1.091 | Cancer |
| Neurocognitive conditions | 0.294 | Neurological Condition |
| Pregnancy, childbirth & puerperium | 0.789 |  |
| COPD x Age | -0.002 | Lung Disease x Age |
| Asthma x Age | -0.015 | Asthma x Age |
| Obesity x Age | -0.004 | Severe Obesity x Age |
| Diabetes x Age | 0.000 | Diabetes x Age |
| Hypertension x Age | 0.005 | Hypertension x Age |
| Congestive heart failure x Age | -0.007 | Heart Disease x Age |
| Myocardial infarction x Age | 0.003 | Heart Attack x Age |
| Rheumatic heart disease x Age | -0.008 |  |
| Stroke x Age | -0.003 | Stroke x Age |
| Sickle cell/HIV/Transplant x Age | -0.028 |  |
| Chronic kidney disease x Age | -0.008 | Kidney Disease x Age |
| Hemodialysis x Age | -0.018 |  |
| Liver disease x Age | 0.001 |  |
| Pneumonia, acute bronchitis, influenza or other acute respiratory infection x Age | -0.005 |  |
| Cancer x Age | -0.009 | Cancer x Age |
| Neurocognitive conditions x Age | 0.004 | Neurological Condition x Age |
| Pregnancy, childbirth & puerperium x Age | -0.003 |  |

## Appendix C Risk Factors by Subgroup and Differences in VI in Midlife by Two Dimensions of Population Subgroup Membership

**Table C-1.**
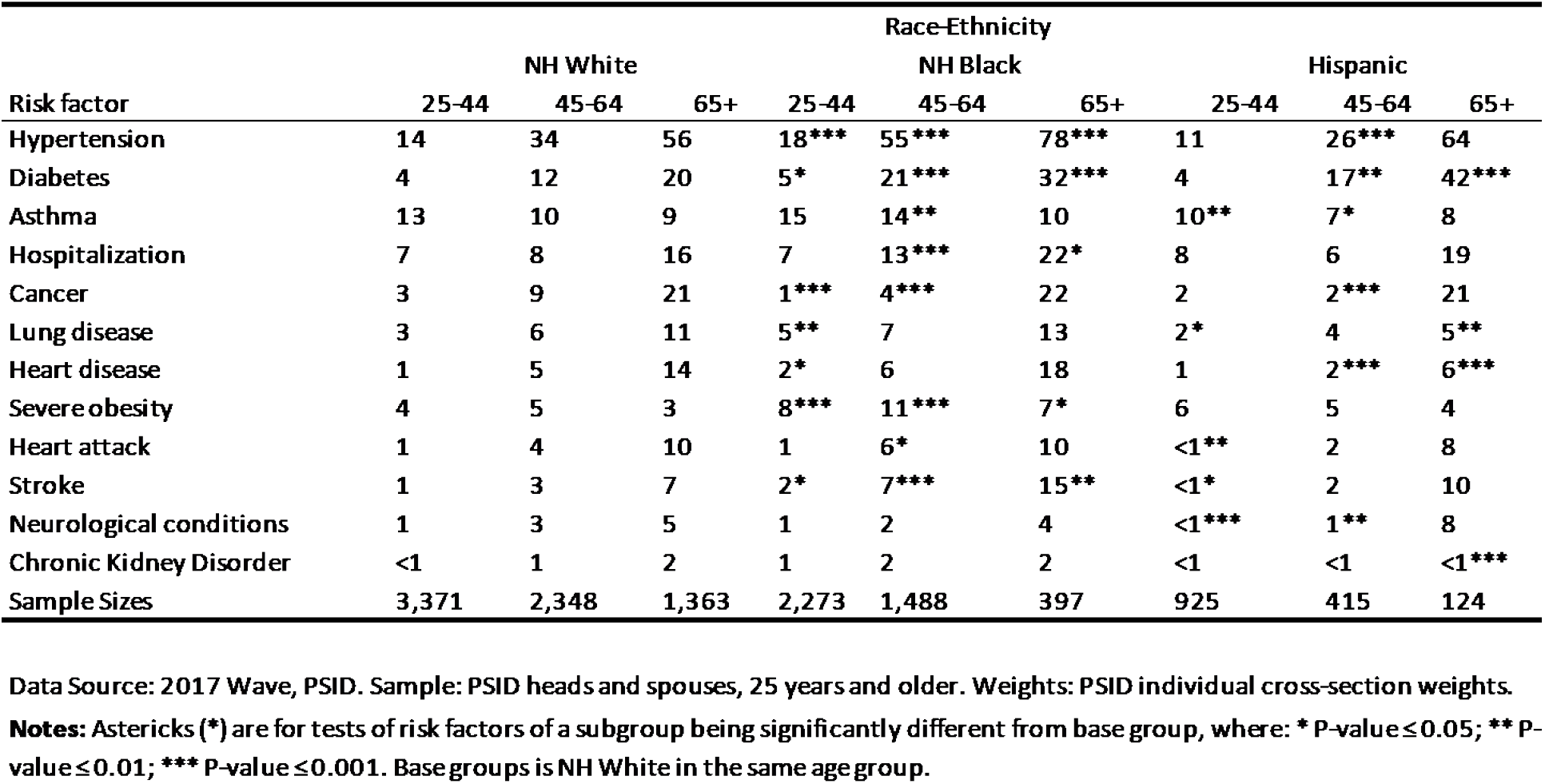
Prevalence (%) of Risk Factors by Race-Ethnicity and Age

**Table C-2.** Prevalence (%) of Risk Factors by Educ and Age

| Risk factor | Education |  |  |  |  |  |
| --- | --- | --- | --- | --- | --- | --- |
|  | HS or less |  |  | BA or more |  |  |
|  | 25-44 | 45-64 | 65+ | 25-44 | 45-64 | 65+ |
| Hypertension | 17*** | 40*** | 62*** | 11 | 28 | 54 |
| Diabetes | 4 | 17*** | 25*** | 4 | 10 | 17 |
| Asthma | 12 | 10 | 11* | 11 | 9 | 8 |
| Hospitalization | 9*** | 11*** | 19*** | 6 | 5 | 13 |
| Cancer | 2 | 6 | 22 | 3 | 7 | 23 |
| Lung disease | 5*** | 8*** | 14*** | 1 | 2 | 6 |
| Heart disease | 1*** | 6*** | 15 | 0 | 3 | 12 |
| Severe obesity | 6*** | 7*** | 4 | 3 | 3 | 3 |
| Heart attack | 1*** | 5*** | 12** | 0 | 1 | 8 |
| Stroke | 1** | 4*** | 9* | 0 | 1 | 6 |
| Neurological conditions | 2*** | 4*** | 7** | 0 | 1 | 4 |
| Chronic Kidney Disorder | <1 | 1 | 1* | 0 | 1 | 3 |
| Sample Sizes | 2,421 | 1,938 | 903 | 2,447 | 1,341 | 618 |

Data Source: 2017 Wave, PSID. Sample: PSID heads and spouses, 25 years and older. Weights: PSID individual cross-section weights.
**Notes:** Asterisks (\*) are for tests of risk factors of a subgroup being significantly different from base group, where: \* P-value $\leq 0.10$ ; \*\* P-value $\leq 0.05$ ; \*\*\* P-value $\leq 0.01$ . Base group is BA or more in the same age group.

**Table C-3.** Prevalence (%) of Risk Factors by Household Income and Age

| Risk factor | Household Income |  |  |  |  |  |
| --- | --- | --- | --- | --- | --- | --- |
|  | Bottom Quartile |  |  | Top Quartile |  |  |
|  | 25-44 | 45-64 | 65+ | 25-44 | 45-64 | 65+ |
| Hypertension | 18*** | 50*** | 67*** | 13 | 28 | 46 |
| Diabetes | 6*** | 21*** | 26*** | 3 | 9 | 14 |
| Asthma | 16*** | 19*** | 11 | 10 | 8 | 8 |
| Hospitalization | 13*** | 18*** | 22*** | 5 | 5 | 12 |
| Cancer | 3 | 8 | 19 | 3 | 8 | 21 |
| Lung disease | 5*** | 17*** | 13*** | 1 | 2 | 6 |
| Heart disease | 3*** | 10*** | 18*** | 0 | 2 | 10 |
| Severe obesity | 7*** | 9*** | 5*** | 3 | 3 | 2 |
| Heart attack | 1 | 6*** | 13** | 1 | 2 | 8 |
| Stroke | 2** | 7*** | 11*** | 0 | 2 | 5 |
| Neurological conditions | 4*** | 8*** | 8*** | 0 | 0 | 1 |
| Chronic Kidney Disorder | <1 | 2** | 1 | 0 | 0 | 2 |
| Sample Sizes | 1,243 | 797 | 510 | 1,830 | 1,552 | 392 |

Data Source: 2017 Wave, PSID. Sample: PSID heads and spouses, 25 years and older. Weights: PSID individual cross-section weights. Income quartiles are determined across households using family weights.
**Notes:** Asterisks (\*) are for tests of risk factors of a subgroup being significantly different from base group, where: \* P-value $\leq 0.10$ ; \*\* P-value $\leq 0.05$ ; \*\*\* P-value $\leq 0.01$ . Base group is top quartile in the same age group.

**Table C-4.**
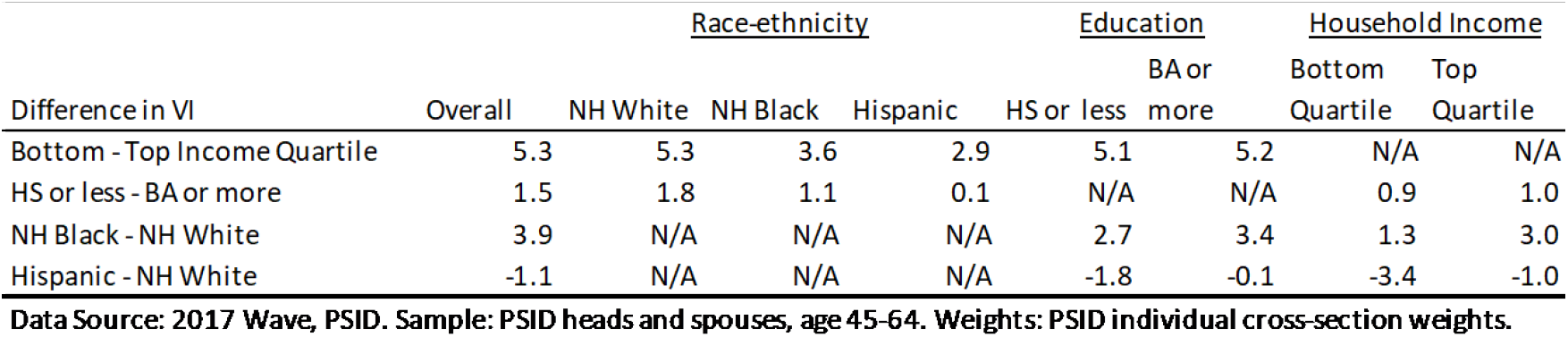
Differences in Median VI Across two Dimensions of Race-Ethnicity, Educational Attainment, and Household Income in Midlife (45-64)

## Appendix D Calculation of *VI* among COVID-19 Hospitalized Patients

To calculate *VI* based on the average conditions of COVID-19 hospitalized patients overall and by age group (18-49, 50-64, 65+), we use the prevalence of preexisting conditions of hospitalized patients reported in Garg et al. (20) and apply these prevalence rates to our calculation of *VI* described in Equation (4) in the text. We match each condition with its corresponding condition in DeCaprio et al. (23) as shown in Appendix Table D-1. For the two demographic factors in DeCaprio et al. (23), age and gender, we apply the median age overall and in each age group from the PSID and the percent of all hospitalized patients who are male (54%) from Garg et al. (20) to each age group. There are several preexisting conditions in Garg et al. (20) that are not in the DeCaprio et al. model and we do not include these in our calculations of *VI*.

Garg et al. (20) on preexisting conditions was only from March 2020 and for 178 hospitalized patients (12% of hospitalized adults), as these data require time-consuming abstractions from hospital medical charts. Though the sample in Garg et al. (20) is small, it has the advantage that preexisting conditions are reported overall and by age group. The incidence of preexisting health conditions for adults is very similar to that data reported for the week ending June 6, 2020, although June data are not reported by age (21).

We have tested using applying the coefficients for neurocognitive conditions instead of stroke as the corresponding condition for neurologic disease and the results are unchanged. We have also tested using sickle cell/HIV/transplant in DeCaprio et al. (2020) as a match for immunosuppresive conditions in Garg et al. (r2020). Again the results are very similar.

**Table D-1.** Correspondence between Garg et al. (2020) and DeCaprio et al. (2020)

| Underlying Conditions Garg et al. (2020) | Corresponding Condition DeCaprio et al. (2020) |
| --- | --- |
| Hypertension | Hypertension |
| Obesity (BMI≥30) | Severe Obesity (BMI≥40) |
| Chronic metabolic disease | No exact match (excluded) |
| Diabetes mellitus | Diabetes |
| Chronic lung disease |  |
| Asthma | Asthma |
| Chronic obstructive pulmonary disease | (COPD) or emphysema, cystic fibrosis, or chronic bronchitis |
| Cardiovascular disease (not hypertension) |  |
| Coronary artery disease | Rheumatic and other heart disease |
| Congestive heart failure | Congestive heart failure |
| Neurologic disease | Stroke |
| Renal disease | Chronic kidney disease |
| Immunosuppressive condition | No exact match (excluded) |
| Gastrointestinal/Liver disease | Liver disease |
| Blood disorder | No exact match (excluded) |
| Rheumatologic/Autoimmune disease | No exact match (excluded) |

**Notes:** Pregnancy is excluded because we do not use it in the PSID estimation. Garg et al. (2020) reports that 30% of obese patients are severely obese and we apply the percentage uniformly across age groups. We treat all reports of chronic lung disease that are not asthma as being included in COPD, emphysema, cystic fibrosis, or chronic bronchitis. We treat all reports of cardiovascular disease (not hypertension) that are not congestive heart disease as coronary artery disease. We have tested using applying the coefficients for neurocognitive conditions instead of stroke as the corresponding condition for neurologic disease and the results are unchanged. We have also tested using sickle cell/HIV/transplant in DeCaprio et al. (2020) as a match for immunosuppressive conditions in Garg et al. (2020). Again the results are very similar.

## Footnotes

1 PSID defines a cohabiting partner is one who has lived with the head for one year or more. Since 2017, the PSID has referred to heads as “reference person 1” and spouses and cohabiters as “reference person 2.” For ease in comparison to past work, we continue to use “head” and “spouse.”

2 There are 13,831 heads and spouses age 25 and older. We dropped 681 due to missing data, 302 due to missing data on race-ethnicity, education, or income and a further 379 due to missing data about health conditions, producing a sample of 13,150.

3 The lowest quartile consisted of those households with reported incomes between $0 and $29,096; the second quartile, $29,096 and $56,760; the third quartile, $56,760 and $101,820; and the top quartile, incomes greater than $101,820. The measure is based on reports about the income received in 2016 and is reported in 2016 dollars.

4 There are slight differences between the PSID and CPS samples for the distributions of educational attainment, presumably because of slightly different questions and their wording.

5 At https://closedloop.ai/c19index/ one can calculate values of what they refer to as the “C-19index” based on risk factors, hospitalization experience and age. The Survey Model that we use is similar, but not exactly the same, as the web model is trained on additional information that we do not use, e.g., zip-code.

6 Similar patterns emerge when using means and medians.

7 The first two columns of Table B-1 are taken from Table 4 of DeCaprio et al. (2020) for their Survey Model. CCSR codes are provided for each of the conditions listed in the model.

## Notes

### Competing Interest Statement

The authors have declared no competing interest.

### Clinical Trial

This study analyzes secondary data.

### Author Declarations

Research covered by Protocol 2017-0566 (C0757) through the the Duke University Campus Institutional Review Board.

